# Prevalence of the major thyroid cancer-associated syndromes in the United States

**DOI:** 10.1101/2024.12.01.24318259

**Authors:** Samantha L. White, Taylor Jamil, Caitlin Bell, Lauren Fishbein, Bryan R. Haugen, Christopher R. Gignoux, Nikita Pozdeyev

## Abstract

**Importance:** A subset of thyroid cancers develops in a setting of a known hereditary cancer-associated syndrome. Understanding the population prevalence of thyroid cancer-associated syndromes is important to guide germline genetic testing and clinical management.

**Objective:** To estimate the prevalence of the major thyroid cancer-associated syndromes in the United States using the All of Us Research Program (AoU) data.

**Design:** In this cohort study, we identified pathogenic and likely pathogenic (P/LP) variants from the ClinVar database in 245,394 AoU biobank participants. We performed a logistic regression analysis of the association of ClinVar P/LP variants with thyroid cancer. P/LP variants in the genes of interest were manually curated to ensure match and pathogenicity status. We calculated the prevalence of thyroid cancer-associated syndromes defined by the presence of P/LP variants.

**Results:** Using logistic regression, we found that three hereditary syndromes, multiple endocrine neoplasia type 2 (MEN2, *RET* gene, p = 3.23e-20), PTEN hamartoma syndrome (PHPS, *PTEN* gene, p = 2.59e-15), and familial adenomatous polyposis type 1 (FAP, *APC* gene, p = 2.73e-10) were significantly associated with thyroid cancer. All these syndromes were previously reported to increase the risk of thyroid cancer. The prevalence of thyroid cancer-associated syndromes in the AoU was 1:2,172 (113 cases), 1:8,764 (28 cases), and 1:8,461 (29 cases) for MEN2, PHPS, and FAP, respectively. Most carriers of P/LP variants were not diagnosed with the features of the syndromes, including thyroid cancer, pheochromocytoma, or primary hyperparathyroidism. Three pathogenic *RET* variants that cause two amino acid substitutions, V804M and V804L, constitute 65% of all MEN2 variants in the AoU, and none of these carriers were diagnosed with thyroid cancer.

**Conclusions and Relevance:** The prevalence of MEN2 and PHPS is ∼10-20 times higher than it is currently estimated for the general population (1:35,000 – 1,50,000 for MEN2 and 1:200,000-1:250,000 for PHTS). Most affected individuals are not diagnosed with thyroid cancer. These results further refine our understanding of the prevalence of hereditary syndromic thyroid cancers and may change the clinical approach to patients with moderate-risk *RET* mutations (such as V804M and V804L), potentially emphasizing active surveillance over prophylactic thyroidectomy.

**Key Points:** 

**Question:** What is the prevalence of the major thyroid cancer-associated syndromes in the United States?

**Findings:** In this cohort study that includes 245,394 genotyped participants in the All of Us Research Program (AoU), the prevalence was 1:2,172 for multiple endocrine neoplasia type 2 (MEN2) and 1:8,764 for PTEN hamartoma syndrome (PHTS).

**Meaning:** The prevalence of MEN2 and PHPS is ∼10-20 times higher than currently estimated for the general population.

## Background

Genetic effects are estimated to contribute up to 53% to the susceptibility to thyroid cancer in family studies,^1,2^ making it one of the most heritable common cancers.^1,3^ Despite the strong contribution of genetic predisposition to thyroid cancer risk, germline genetic risk assessments are not frequently used for clinical care, which is a missed opportunity to improve thyroid cancer screening, detection, and treatment.

Some thyroid cancers develop in the setting of genetic cancer-associated syndromes. Medullary thyroid carcinoma (MTC) is a component of multiple endocrine neoplasia types 2 and 3 (MEN, *RET* gene).^4^ Follicular cell-derived thyroid cancers occur in PTEN hamartoma syndrome (PHTS or Cowden syndrome, *PTEN* gene),^5^ familial adenomatous polyposis type 1 (FAP or Garner syndrome, *APC* gene),^6^ Carney complex (*PRKAR1A* gene),^7^ Li-Fraumeni syndrome (*TP53* gene)^8^ and Werner syndrome (*WRN* gene)^9^. These thyroid cancer-associated syndromes are thought to be rare, with the population prevalence ranging from 1:7000 for FAP^6^ to 1:600,000-4,000,000 for MEN3 (formerly known as MEN2B).^10^

Due to the growth of large biobanks such as the All of Us Research Program (AoU)^11^ and Colorado Center for Personalized Medicine (CCPM)^12^, more people in the United States get their genomes sequenced irrespectively of the clinical indication. This leads to the incidental discovery of actionable variants in the reportable genes on the American College of Medical Genetics and Genomics (ACMG) list.^13^ Based on our clinical experience evaluating patients with P/LP mutations discovered on genotyping by the CCPM, we hypothesized that thyroid cancer-associated syndromes are more prevalent in the population than previously estimated. We believe that many individuals in the population at risk of developing syndromic thyroid cancer have not yet been discovered by the healthcare system, suggesting higher prevalence and lower penetrance than previously estimated.

Large population-scale studies genotyping hundreds of thousands of individuals and linking genetic and clinical data, such as the AoU,^14^ permit an unbiased estimate of hereditary disease prevalence by exploring known pathogenic variants.

## Methods

All analyses were performed on the AoU Controlled Tier Dataset v7 (n = 245,394) using clinical, survey, and short-read whole genome sequencing data. In compliance with the AoU Data User Code of Conduct, the computer code written in *R, Python* and *SQL* (https://github.com/pozdeyevlab/clinvar-tc) was executed on the AoU Research Workbench (workspace “Thyroid Cancer Genetics v7”), and data corresponding to fewer than 20 participants was obscured before publication.

### Phenotype definitions

Thyroid cancer phenotype was defined by the presence of at least one condition code for the “Carcinoma in situ of thyroid gland” [Observational Medical Outcomes Partnership (OMOP) Common Data Model concept ID 4241917], “Malignant tumor of thyroid gland” (4178976), “Follicular thyroid carcinoma” (4111010), “Hurthle cell carcinoma of thyroid” (4307263), and “Primary differentiated carcinoma of thyroid gland” (36676291). In addition, we used survey codes: “Including yourself, who in your family has had thyroid cancer? Select all that apply. – Self”, “Are you still seeing a doctor or health care provider for thyroid cancer? – Yes”, “About how old were you when you were first told you had thyroid cancer?” (any age), “Are you currently prescribed medications and/or receiving treatment for thyroid cancer? – Yes”. Pheochromocytoma and primary hyperparathyroidism were defined using OMOP concept IDs 136934 and 4118993, respectively. To estimate the prevalence of clinical disease, we used only AoU participants with either electronic health records or survey data available (n = 237,708).

Age at thyroid cancer diagnosis was extracted from the ‘condition_occurrence’ table in the AoU Curated Data Repository and defined as age at the first occurrence of the thyroid cancer ICD-9-CM or ICD-10-CM billing code (193 and C73, respectively). Age is presented as a mean ± standard deviation.

### Pathogenic variants

Genetic variants in *RET, PTEN, APC, DICER1, POT1, MSH2, MSH6, MLH1, PMS2, NF1, NF2, SDHA, SDHB, SDHC, SDHD, MUTYH, STK11*, and other genes for genetic syndromes with an autosomal dominant mode of inheritance were downloaded from the ClinVar database,^15^ September 2023 release. To avoid overreporting cancer-associated syndromes, we extracted only single nucleotide polymorphisms and indels reported as pathogenic or likely pathogenic (P/LP). P/LP variants identified in the AoU dataset were manually curated to ensure that a variant record with matching position, reference, and alternate alleles exists in the ClinVar and the pathogenicity classification is P/LP as of October 2024.

### Logistic regression analysis

We tested ClinVar genetic syndromes that have at least one P/LP variant in the dataset for the association with thyroid cancer using multiple logistic regression analysis. The regression model was adjusted for sex, age, and 16 genetic principal components. P-values were reported by syndrome, and the significance threshold was adjusted using Bonferroni correction.

## Results

We found 2,097 genotyped patients with a history of thyroid cancer in the AoU version 7 dataset, which corresponds to a thyroid cancer population prevalence of 0.88%.

### Genetic syndromes associated with thyroid cancer

Logistic regression analysis of genetic syndromes reported in the ClinVar database identified three diseases significantly associated with thyroid cancer: multiple endocrine neoplasia type 2 (*RET*), PTEN hamartoma syndrome (*PTEN*), and familial adenomatous polyposis (*APC*) (**Table 1**). All three syndromes are known to cause thyroid cancer.^4-6^

**Table 1.** Hereditary syndromes significantly associated with thyroid cancer in the All of Us Research Program Biobank.

| Hereditary syndrome | Gene | p-value<sup>§</sup> |
| --- | --- | --- |
| Multiple endocrine neoplasia, type 2 | *RET* | 3.23e-20 |
| PTEN hamartoma tumor syndrome | *PTEN* | 2.59e-15 |
| Familial adenomatous polyposis 1 | *APC* | 2.73e-10 |

<sup>§</sup> p-values significant at a Bonferroni-corrected p-value threshold ( $\leq 3.36\text{e-}5$ ) are shown.

### RET

There are 113 carriers of P/LP variants in the *RET* gene in the entire genotyped AoU version 7 cohort, resulting in a population prevalence of MEN2 of 1:2,172 (113:245,394) or 0.046%. We did not find *RET* variants for MEN type 3 syndrome. Ninety-one of P/LP *RET* variants are described in the revised American Thyroid Association (ATA) MTC guidelines.^4^ Using a more strict definition of MEN2 by only including *RET* variants listed in the ATA guidelines, the prevalence of the disease in AoU is 1:2,697 (91:245,394).

All carriers of P/LP *RET* variants are heterozygous. Almost all P/LP *RET* variants are classified as moderate risk per the ATA.^4^

The most common pathogenic variation in RET protein in AoU participants occurs at the amino acid 804 (V804M and V804L) caused by three pathogenic variants, chr10:43119548:G:A (ClinVar variation ID 37102), chr10:43119548:G:C (ClinVar variation ID 38613), and chr10:43119548:G:T (ClinVar variation ID 13946). Altogether, these three variants account for 73 MEN2 cases or 65% of all MEN2 cases in the AoU dataset. Few (≤20) AoU participants with MEN2 were diagnosed with any clinical features of MEN2 including thyroid cancer, pheochromocytoma, or primary hyperparathyroidism. None of the RET V804M or V804L mutation carriers were diagnosed with thyroid cancer.

The MEN2 prevalence was not different by sex at birth (Table 2, χ^2^, p = 0.14). For those *RET* P/LPV carriers who had thyroid cancer, the age at thyroid cancer diagnosis was 55.8 ± 17.8 years.

**Table 2.** Age and counts for AoU participants with thyroid cancer-associated syndromes.

| Cohort | Mean (SD) age, years |  | N<sup>b</sup> |  |  |
| --- | --- | --- | --- | --- | --- |
|  | At the time of analysis<sup>a</sup> | At thyroid cancer diagnosis | All | Female<sup>c</sup> | Male<sup>c</sup> |
| All | 57 (17) |  | 245,388 | 145,563 | 94,756 |
| MEN2 | 56 (18) |  | 113 | 76 | 36 |
| MEN2 diagnosed with thyroid cancer | 64 (21) | 56 (20) |  |  |  |
| MEN2 not diagnosed with thyroid cancer | 55 (17) |  |  |  |  |
| PTEN hamartoma syndrome | 48 (14) |  | 28 |  |  |
| PTEN hamartoma syndrome diagnosed with thyroid cancer | 40 (14) | 28 (16) |  |  |  |
| FAP | 47 ± 14 |  | 29 |  |  |
| FAP diagnosed with thyroid cancer | 50 ± 11 | 37 ± 20 |  |  |  |

<sup>a</sup> - age as of October 2024 is shown.
<sup>b</sup> - only N >20 are shown in compliance with the AoU Data User Code of Conduct
<sup>c</sup> - sex at birth when known

### PTEN

There are 28 P/LP variants in the *PTEN* gene in the AoU, resulting in a population prevalence of PTEN hamartoma syndrome of 1:8,764 (28:245,394). All carriers are heterozygotes. Only a small fraction of individuals with P/LP variants in the *PTEN* gene were diagnosed with thyroid cancer. The age at thyroid cancer diagnosis was 28 ± 16 years.

### APC

There are 29 P/LP variants in the *APC* gene in the AoU, resulting in a population prevalence of FAP of 1:8,461 (29:245,394). All carriers are heterozygotes. Only a few individuals with P/LP variants in the *APC* gene were diagnosed with thyroid cancer. The age at thyroid cancer diagnosis was 37 ± 20 years.

### Other genes

We found thyroid cancer patients carrying P/LP variants in *BRCA1* and *BRCA2* genes (breast-ovarian cancer susceptibility syndrome) *SDHB, SDHC, SDHD* (hereditary paraganglioma-pheochromocytoma syndrome), *MSH6, MLH1, PMS2* (Lynch syndrome) and *NF1* (neurofibromatosis type 1) genes. It is unclear if these variants are causal for thyroid cancer. Overall, one in fifty-three AoU participants with a history of thyroid cancer carry a known P/LP variant in an actional cancer-predisposing gene on the ACMG list.^13^

None of the patients with thyroid cancer had P/LP variants in *TP53* (Li-Fraumeni syndrome), *DICER1* (DICER1 syndrome), *STK11* (Peutz-Jeghers syndrome) *MUTYH* (MUTYH-associated polyposis), *NF2* (neurofibromatosis type 2) or *POT1* (POT1 tumor predisposition syndrome).

## Discussion

In this study, we report that the prevalence of thyroid cancer-associated syndromes, based on the presence of P/LP variants in known susceptibility genes, is much higher in the AoU than previously reported for the general population. For example, the prevalence of MEN2 syndrome in the United States is thought to be between 1:35,000 and 1:50,000,^10,16^ which is remarkably less than our estimate of ∼1:2,200 – 1:2,800. This discrepancy could be due to clinically missed disease or due to lower-than-expected penetrance of disease. Low penetrance is suggested by the observation that most carriers of P/LP *RET* variants in our study were not diagnosed with thyroid cancer, pheochromocytoma, primary hyperparathyroidism, or MEN2 and, therefore, would not be counted by previous studies.

Age does not fully explain why only a fraction of RET P/LP variant carriers were diagnosed with thyroid cancer. The mean age of people with MEN2 in AoU is 56 ± 18, and age at thyroid cancer diagnosis was not significantly different from age at the time of analysis for RET P/LP carriers that do not have thyroid cancer diagnosis (t-test, p =0.95).

Most P/LP *RET* variants in the AoU are considered moderate risk per the ATA guidelines^4^ and may have low penetrance or cause mild subclinical disease. This hypothesis is supported by the earlier reports that RET V804M and V804L mutations (contributing to 65% of MEN2 cases in the AoU) result in a mild phenotype with late penetrance, C-cell hyperplasia, or indolent MTC.^17^ Most carriers of RET V804M and V804L mutations develop MTC after the age of 50 years.^18-20^

Aggressive MTC in RET V804M and V804L mutation carriers presenting early with lymph node metastases has been described.^21-25^ Consequently, ATA guidelines^4^ suggest that prophylactic thyroidectomy can be performed to prevent morbidity. Our data suggests that a conservative screening and surveillance approach to patients with moderate-risk RET mutations may be appropriate, as none of the carriers of RET V804M or V804L moderate-risk variants were diagnosed with thyroid cancer. Alternatively, prophylactic thyroidectomy can be postponed until later in life (≥ 50 years). Further supporting conservative management, occult MTC was found in 0.14% of 7,897 autopsies of patients who died from other causes.^26^ This data confirms that there is a large pool of people in the population who have subclinical disease that may not require treatment. A more restrained approach to the management of moderate-risk RET mutations is expected to reduce complications from thyroidectomies and the incidence of iatrogenic hypothyroidism at a young age.

The reason why RET V804M and V804L are the most common RET variants seen in the AoU is not known. It is possible that the low penetrance and indolent phenotype allow these variants to escape negative genetic selection. Alternatively, a founder effect like the one described for RET C611Y in the Danish population^27^ may play a role.

PTHS is a genetic syndrome associated with follicular cell-derived thyroid cancer such as papillary and follicular thyroid cancer. Similar to MEN2, we found a much higher prevalence of PTHS in the AoU than the previously reported prevalence of 1:200,000 and 1:250,000.^28,29^ The lifetime risk of thyroid cancer in *PTEN* mutation carriers has been reported to be 3% to 10%^30^ matching our findings in AoU. The prevalence of FAP associated with the cribriform-morular variant of papillary thyroid cancer^31^ in the AoU (∼ 1:8,500) is similar to previous estimates.^6^

Age at thyroid cancer diagnosis was significantly higher for MEN2 (56 ± 20 years, Table 2) than for PTHS (28 ± 16 years, t-test, p=0.03) but not FAP (37 ± 20 years, t-test, p=0.24). This may be explained by a greater pathogenicity of some *PTEN* P/LP variants. Alternatively, younger age at thyroid cancer diagnosis could be due to thyroid cancer screening of PTHS patients diagnosed early due to extrathyroidal manifestations.

We found that 1 in 53 patients diagnosed with thyroid cancer carry an actionable P/LP variant leading to cancer-associated syndrome. This relatively high occurrence highlights the importance of careful collection of family history in patients with thyroid cancer and consideration of the referral for genetic counseling and germline genetic testing of high-risk individuals.

This study has limitations. The AoU participants are not recruited via probability sampling of the United States population. However, the population prevalence of thyroid cancer in the AoU (0.88%) is similar to the population-based data from the American Cancer Society^32^ indicating that AoU is not biased towards a higher prevalence of thyroid cancer. Most of the clinical data was extracted from electronic health records and may be incomplete, resulting in the underestimation of the proportion of P/LP variant carriers diagnosed with thyroid cancer. It is also not known how many of the carriers of P/LP variants leading to thyroid cancer-associated syndromes will be diagnosed with clinically significant malignancy later in life.

In summary, the incidental findings of P/LP variants in *RET, PTEN*, and other thyroid cancer-associated genes will become more frequent due to the recruitment of people from the general population into large biobanks such as AoU and the increased availability of clinical genetic testing. We found that most of these individuals have not been diagnosed with thyroid cancer. It is likely that active surveillance with neck ultrasound and serum calcitonin for most of the carriers of moderate-risk mutations in RET is appropriate, and the risk of overtreatment is high.

## Data Availability

All data produced in the present work are contained in the manuscript

## Acknowledgments

We gratefully acknowledge All of Us participants for their contributions, without whom this research would not have been possible. We also thank the National Institutes of Health’s All of Us Research Program for making available the participant data examined in this study.

